# The utility of infectious disease modelling in informing policy for outbreak response: a scoping review

**DOI:** 10.1101/2025.03.04.25323088

**Authors:** D Rao, A Tanveer, EN Iftekhar, SA Müller, K Sherratt, K Röbl, P Carrillo-Bustamante, K Heldt, J Fitzner, J Hanefeld, S Funk

**Affiliations:** Centre for International Health Protection, Robert Koch Institute, Berlin, Germany; Institute of International Health, Charité—Universitätsmedizin Berlin, Berlin, Germany; Centre for Mathematical Modelling of Infectious Diseases, London School of Hygiene & Tropical Medicine, London, United Kingdom; World Health Organization Hub for Pandemic and Epidemic Intelligence, Berlin, Germany; Department of Global Health and Development, London School of Hygiene & Tropical Medicine, London, United Kingdom

**Author notes:** First authors (D Rao, A Tanveer, EN Iftekhar).

## Abstract

**Background and objectives:** Infectious disease modelling plays a critical role in guiding policy during outbreaks. However, ongoing debates over the utility of these models highlight the need for a deeper understanding of their role in policymaking. In this scoping review we sought to assess how infectious disease modelling informs policy, focusing on challenges and facilitators of translating modelling insights into actionable policies.

**Methods:** We searched the Ovid database to identify modelling studies that included an assessment of utility in informing policy and decision-making from January 2019 onwards. We further identified studies based on expert judgement. Results were analysed descriptively. The study was registered on the Open Science Framework platform.

**Results:** Out of 4007 screened and 12 additionally suggested studies, a total of 33 studies were selected for our review. None of the included articles provided objective assessments of utility but rather reflected subjectively on modelling efforts and highlighted individual key aspects for utility. 27 of the included articles considered the COVID-19 pandemic and 25 of the articles were from high-income countries. Most modelling efforts aimed to forecast outbreaks and evaluate mitigation strategies. Participatory stakeholder engagement and collaboration between academia, policy, and non-governmental organizations were identified as key facilitators of the modelling-to-policy pathway. However, barriers such as data inconsistencies and quality, uncoordinated decision-making, limited funding and misinterpretation of uncertainties hindered effective use of modelling in decision-making.

**Conclusion:** While our review identifies crucial facilitators and barriers for the modelling-to-policy pathway, the lack of rigorous assessments of the utility of modelling for policy highlights the need to systematically evaluate the impact of infectious disease modelling on policy in future.

## INTRODUCTION

### Context: Infectious disease modelling as a tool for outbreak response

Policymakers in public health face complex decisions during infectious disease outbreaks, such as AIDS, Ebola, Mpox and COVID-19. These decisions often involve balancing multiple competing priorities under significant uncertainty, including the timing and scale of interventions, resource allocation, and the social and economic impacts of control measures (e.g. (1)). This complexity underscores the critical need for effective decision-making tools. Mathematical modelling has emerged as an important tool in this context, often referred to as ‘infectious disease modelling’ (e.g. (2)). It supports outbreak response in multiple ways: Modellers can for example estimate key epidemiological parameters, forecast outbreak size and assess the potential impact of different control measures (e.g. (3, 4)). Thereby, modelling translates complex epidemiological data into actionable insights for decision-makers as highlighted by its use during outbreaks of e.g. A/H1N1 influenza, Ebola and COVID-19 (e.g. (3, 4)).

### Context: Challenges in the use of infectious disease modelling for outbreak response

However, there is extensive debate about whether these models could be trusted and used effectively to guide policy choices (e.g. (1)). Especially when there are conflicting results, models face criticism from politicians, the press, experts from other disciplines and the public (e.g. (3)). These tensions sometimes cause potentially avoidable delays in implementing measures and makes modellers apprehensive to further contributing to outbreak response (e.g. (5)). Hence, it can be challenging to utilise modelling successfully for outbreak response.

### Context: Need for a better understanding of the use of infectious disease modelling for outbreak response

These challenges highlight the need for continued efforts in enhancing how the results of infectious disease modelling for outbreak response are used, as recognised by the ‘Lancet Commission on Strengthening the Use of Epidemiological Modeling of Emerging and Pandemic Infectious Diseases’(6).

### Rationale: Need for a scoping review

Previously, a scoping review on evidence use for decision-making during infectious disease outbreaks was conducted that tangentially considers the role of modelling evidence (7). However, as it lacks a detailed perspective on modelling, focuses on Europe and considers records only from before the COVID- 19 pandemic, an updated more comprehensive overview is needed.

To improve the utility of modelling for policy, it is necessary to identify typical characteristics, facilitators and challenges of the modelling to policy pathway. Such mapping is particularly timely given the ongoing need to strengthen the science-policy interface in outbreak response, such as for the ongoing Mpox epidemic.

At the time of writing, there are two other projects similar to ours: First, there is a pre-registered scoping review focused on the question ‘What defined policy-relevant advanced analytics over time during the COVID-19 pandemic in Europe?’ (8). Second, there is a study that was published after we had already conducted and pre-registered our search and screening; it also contains a scoping review focusing on the effective translation of ‘modelled evidence into policy decisions’ in Lower- and Middle-Income Countries in the context of the COVID-19 pandemic (9). We complement this work by conducting a scoping review that is not limited geographically and also considers diseases beyond COVID-19.

### Questions and objectives: What has been the utility of infectious disease modelling in informing policies for outbreak response?

This review examines how mathematical modelling has informed policy decisions during infectious disease outbreaks and epidemics. We intended to investigate what types of infectious disease models have been used and how they have been utilised in policy decision-making, with particular focus on evaluations of how useful models were in informing disease outbreak control or mitigation efforts.

Our analysis addresses several interconnected questions about modelling-to-policy translation: What characterizes modelling-to-policy processes? What are the facilitators and barriers for successful integration of modelling evidence into policy? We examine publications that reflect on or evaluate modelling engagement with policies for outbreak response, including who has studied these questions, in what contexts, and what recommendations emerge for improving modelling-to-policy translation.

## METHODS

A review protocol for this study has previously been published with the Open Science Framework on June 5 2024, with updates on June 6 2024, July 17 2024 and 20 February 2025, and can be accessed at https://osf.io/cd4qa/ (10). The writing of this manuscript followed the PRISMA guidelines for Scoping Reviews (PRISMA-ScR) (11).

### Eligibility criteria

The eligibility criteria are based on the PCC framework (population, concept and context), as suggested by the Joanna Briggs Institute (12). The detailed inclusion and exclusion criteria by categories as well as types of evidence sources are presented in Table 1.

**Table 1:** Eligibility criteria for publications in the review and the rationales for selecting these criteria.

| **<u>Category</u>** | **<u>Criterion</u>** (inclusion: white, exclusion: red) | **<u>Rationale</u>** |
| --- | --- | --- |
| **Population** | Models on humans | We are interested in outbreaks in human populations. |
| **Concept** | Models to inform disease outbreak control or mitigation efforts (including policy making regarding resource management such as hospital management) | We are interested in the role of models in informing policies for outbreak response. Outbreak response entails control and mitigation efforts. Resource management falls under these efforts. |
|  | Publications that reflect on or evaluate their engagement with policies for outbreak response. | We are interested in the role of models in informing policies for outbreak response. |
|  | Models to inform pharmaceutical interventions | The research body on these models is too big for the scope of this review. |
|  | Models on any specific non-pharmaceutical interventions during the COVID-19 pandemic | There are multiple already ongoing and performed reviews and a large evidence base. |
|  | Models with only theoretical link to policy making | We are interested in the role of models in *actually* informing policies for outbreak response. |
|  | New model generation or comparative models |  |
| **Context** | Models in response to acute infectious disease outbreaks | We are interested in the role of models in informing policies for acute infectious disease outbreak response. |
|  | Models on pathogens SARS-CoV-2, measles, Zika, C. diphtheriae, Mpox, HIV, TB, influenza and Ebola | To keep the scope manageable, we focus on some key pathogens only. |
|  | Models on cost-effectiveness | The research body on these models is too big for the scope of this review. |
| **Type of evidence sources** | Publications: peer reviewed | Peer-review indicates a basic quality control of the considered publications. |
|  | All type of studies | We are interested in any type of study that provides information about the role of models in informing policies for outbreak response. |
|  | Languages: English | As our working language was English, we chose to include only English publications in the search. |
|  | Time: last 5 years (January 2019 - May 2024) | We are interested in current practice. Starting in 2019 also allows us to include the vast literature related to the COVID-19 pandemic. |
|  | Reviews | We want to understand the specific modelling and policy context for each case study. |

### Search strategy

To define the search components, we adapted the CoCoPop scheme (Condition, context, population) to CoCoPop +E to include the evaluation component (13). We searched the database Ovid (Medline, Embase) on June 5 2024 using the search strategy depicted in Supplementary Figure 1. The rationales for the details of the search are listed in Supplementary Table 1.

Following the search, all identified citations were collected and uploaded into EndNote X7 (Clarivate Analytics, PA, USA) and duplicates were removed. Titles and abstracts were screened by the research team for assessment against the eligibility criteria for the review. Additional literature was added through expert judgement in two ways: (a) The senior authors of this review suggested relevant additional publications and (b) the selected literature of a similar scoping review (9) was considered. These additional publications were also checked against the eligibility criteria and included/excluded accordingly.

### Article selection

The full text of selected citations was assessed in detail against the eligibility criteria by the research team. Two reviewers, AT and DR, independently screened all articles for inclusion. Disagreements were resolved by additional reviewers.

### Data extraction

A data extraction table (via Google Sheets) was developed and updated in an iterative way, resulting in five broad and 21 sub categories including their definitions (Table 2). The table updates comprised clarifications of definitions as well as re-conceptionalisations and re-clustering of categories to achieve better coherence.

**Table 2:**
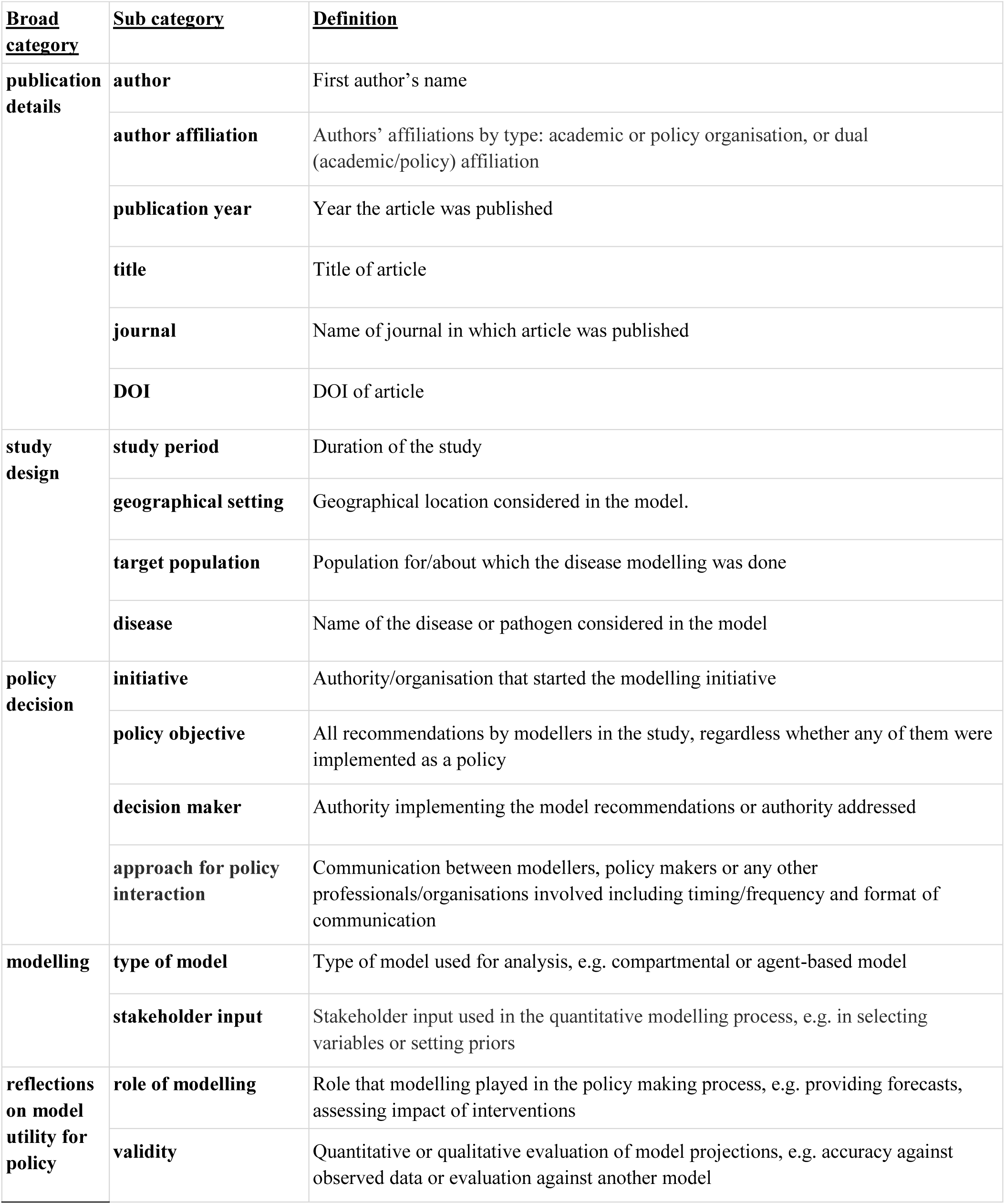
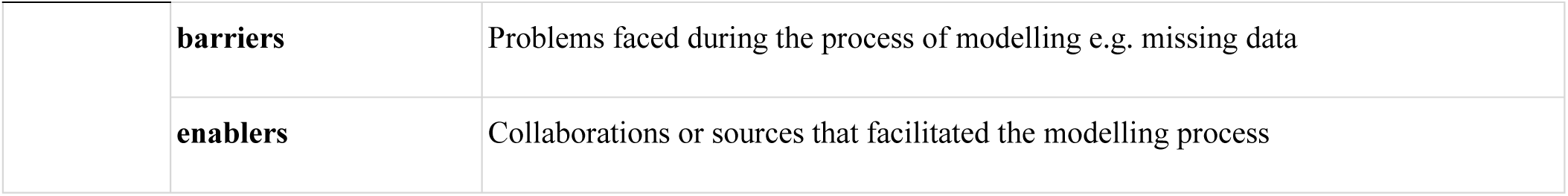
Broad and sub categories for the data extraction along with their definitions.

| **<u>Broad category</u>** | **<u>Sub category</u>** | **<u>Definition</u>** |
| --- | --- | --- |
| **publication details** | **author** | First author's name |
|  | **author affiliation** | Authors' affiliations by type: academic or policy organisation, or dual (academic/policy) affiliation |
|  | **publication year** | Year the article was published |
|  | **title** | Title of article |
|  | **journal** | Name of journal in which article was published |
|  | **DOI** | DOI of article |
| **study design** | **study period** | Duration of the study |
|  | **geographical setting** | Geographical location considered in the model. |
|  | **target population** | Population for/about which the disease modelling was done |
|  | **disease** | Name of the disease or pathogen considered in the model |
| **policy decision** | **initiative** | Authority/organisation that started the modelling initiative |
|  | **policy objective** | All recommendations by modellers in the study, regardless whether any of them were implemented as a policy |
|  | **decision maker** | Authority implementing the model recommendations or authority addressed |
|  | **approach for policy interaction** | Communication between modellers, policy makers or any other professionals/organisations involved including timing/frequency and format of communication |
| **modelling** | **type of model** | Type of model used for analysis, e.g. compartmental or agent-based model |
|  | **stakeholder input** | Stakeholder input used in the quantitative modelling process, e.g. in selecting variables or setting priors |
| **reflections on model utility for policy** | **role of modelling** | Role that modelling played in the policy making process, e.g. providing forecasts, assessing impact of interventions |
|  | **validity** | Quantitative or qualitative evaluation of model projections, e.g. accuracy against observed data or evaluation against another model |
|  | **barriers** | Problems faced during the process of modelling e.g. missing data |
|  | **enablers** | Collaborations or sources that facilitated the modelling process |

Data from each included publication was extracted independently by two reviewers, respectively. The involved reviewers were AT, DR, and ENI. In case of immediate agreement, the wording of the first reviewer was chosen. Complementary data was added, where necessary. Any arising disagreements were discussed and resolved between the two reviewers. Where the data extraction for certain publications was deemed difficult by the reviewers, help was sought from additional co-authors.

### Synthesis of results

The purpose of this scoping review is to map the research done on the utility of infectious disease modelling in informing policy for outbreak response, as well as to identify any existing gaps in knowledge on this topic. As we did not find objective assessments of utility to include into our review but subjective reflections by practitioners, we instead summarised aspects of the modelling to policy process that are claimed to be essential for the utility by the authors of the included publication. We performed a descriptive qualitative content analysis analysis and synthesis of the data extraction results along the topics that we chose in our data extraction table.

## RESULTS

### Article selection

Out of the 4007 articles identified through database search via 3 databases 3965 were screened for inclusion after removal of duplicates (Fig 1). Out of the title and abstract screened articles, the full texts of 42 articles were screened against the eligibility criteria and 26 of them were included in the review. Full texts of additional 12 articles suggested based on expert judgement were also screened against the criteria and 7 of them were included in the review. Data was extracted from all 33 included articles (Supplementary Material).

**Fig 1:**
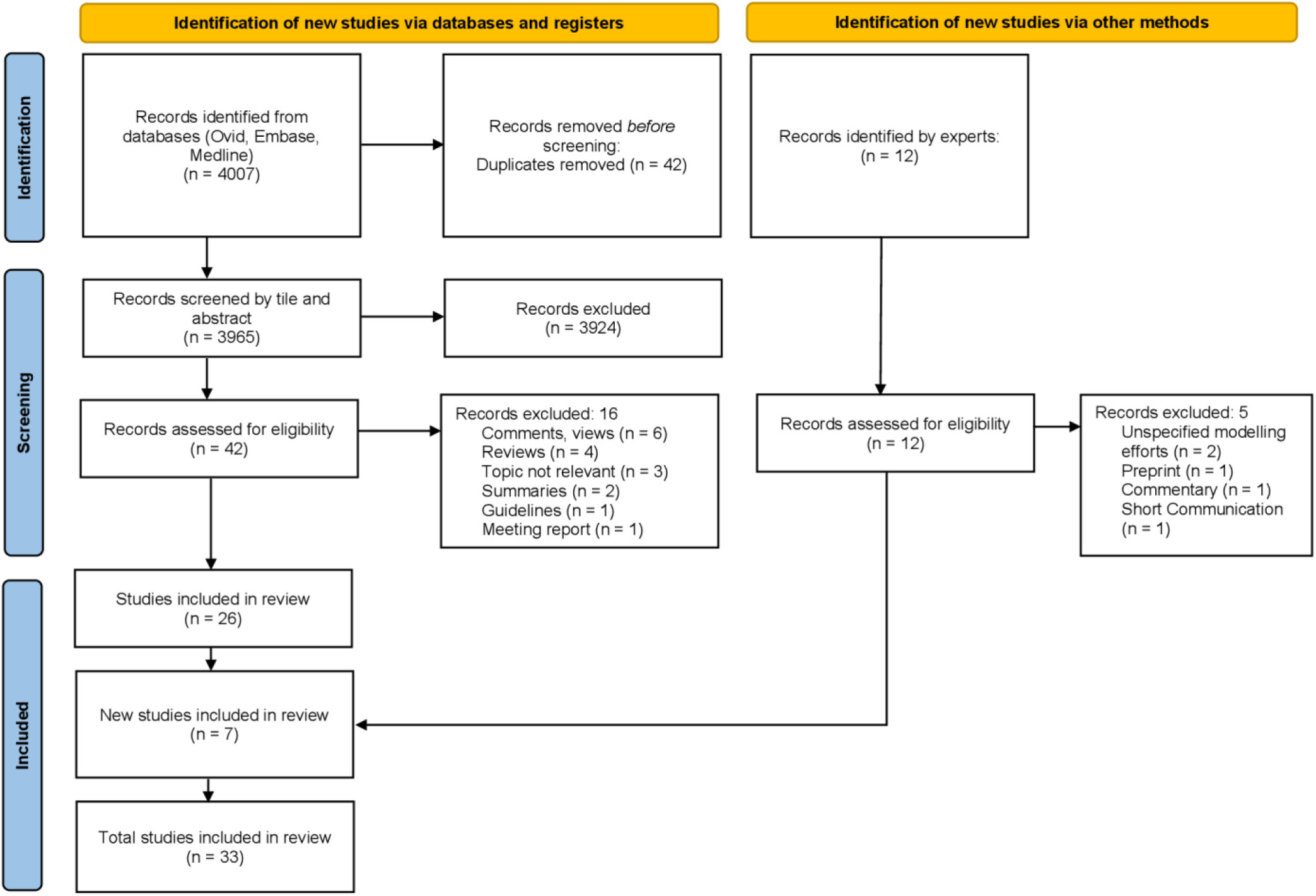
Flow diagram of the article selection process.

### Summary of extracted data

The full extracted data are presented in the data extraction matrix (Supplementary Material).

No study provided objective assessments of utility. 31 of the 33 included articles are descriptions and reflections by modellers about their own modelling efforts where they highlight aspects they deemed relevant for the utility of their work in informing policy for outbreak response. Only in two articles, the perspectives of decision makers are portrayed (14, 15).

The included literature represents all continents (Fig 2). However, most of the studies come from High Income Countries, with multiple articles on modelling efforts in the USA (13 articles) and UK (5 articles). Most Low-and-Middle Income Countries (LMICs) that are highlighted in the graph are considered through the included articles related to the same modelling consortium (‘CoMo Consortium’) (16, 17). These articles were included based on the study by Owek et al. (9).

**Fig 2:**
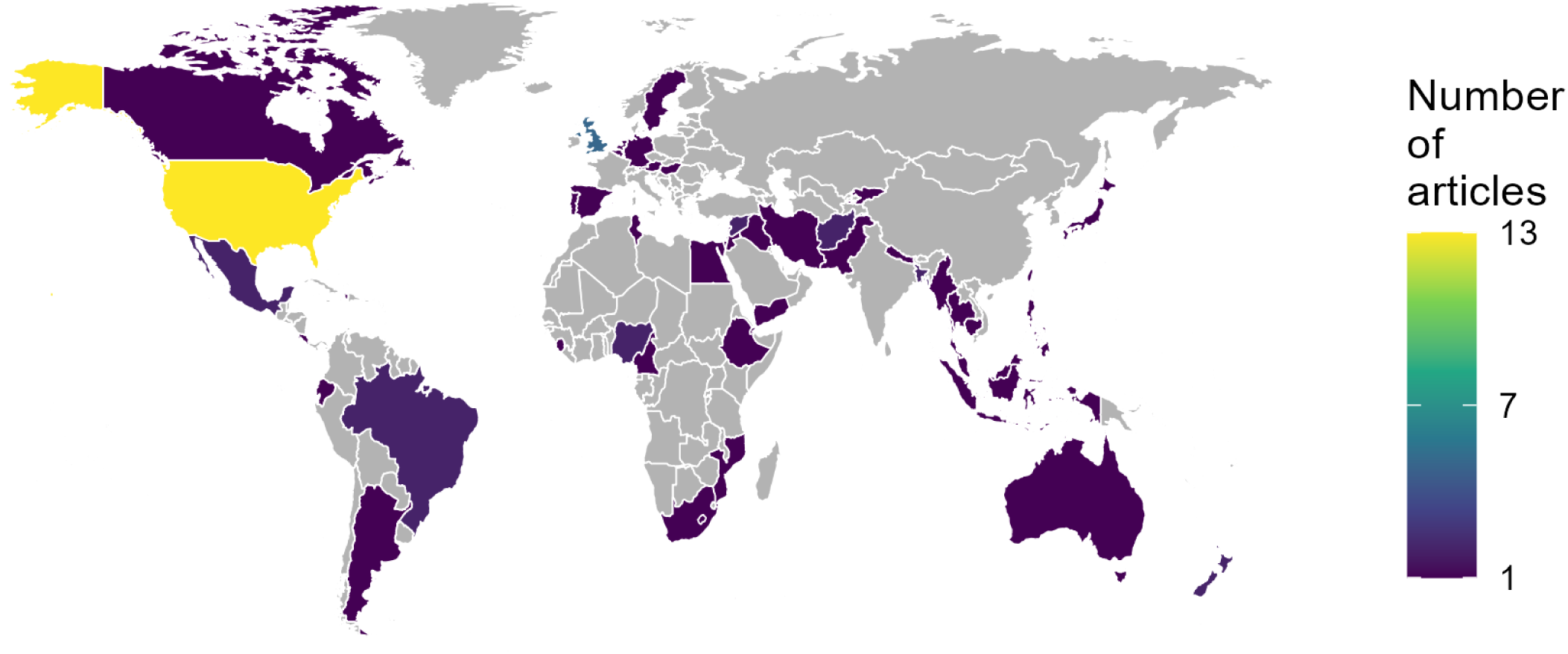
Many countries are represented. World map highlighting the countries that were considered in the included articles. Although all continents are represented, the majority of articles consider the Global North. Note that some articles consider multiple countries; two of these articles account for most of the countries of the Global South in this figure (16, 17).

Out of the 33 articles included, 27 had models on COVID-19, one combined modelling of COVID-19 and Influenza (18) with two models on Influenza alone (19, 20), one on Diphtheria (21), one on HIV (22) and one article considered models on various infectious diseases including Influenza, Pertussis, Tuberculosis and others (23). Our data extraction yielded no key differences between modelling efforts about COVID- 19 and other diseases.

Visualisations of the considered diseases, publication years and considered population groups can be found in the Supplementary Information (Supplementary Figures 2, 3 and 4).

### Authors’ affiliations and modelling initiatives

Out of the 33 included articles, 17 articles were by authors with solely academic affiliations while the remaining 16 articles were authored by both those with academic and policy affiliations.

Infectious disease modelling efforts were initiated at various levels (Table 3). While most modelling efforts were conceptualised and started either by scientists working in academia or by officials at decision- making authorities, some initiatives were also seen as a collaborative work between academia and local, regional or national decision-making authorities. All modelling initiatives were found to be in line with the modelling infrastructures defined in a previous global analysis on the use of epidemiological modelling in health crises, i.e. “one modelling team; multiple small teams functioning as one; a consortium consisting of multiple teams and multiple models led by a modelling committee; and finally, teams working in isolation feeding independently into government” (2).

**Table 3:** Disease modelling efforts were initiated by various actors. . These included policy actors and decision makers but also the modellers themselves.

| Modelling initiatives | Example(s) |
| --- | --- |
| Academia | • Scientists working pro bono for national COVID-19 response, Israel (24)<br>• CoMo Consortium, LMICs (17) |
| Collaboration between academia and policy | • Collaboration between the Ministry of Health, Pan American Health Organisation and scientists from academia; Costa Rica (25)<br>• South African COVID-19 Modelling Consortium, a collaborative effort between academic and government structures (26) |
| Policy and decision-making authority | • Austrian governmental crisis unit, Ministry of Health (27);<br>• UK Health Security Agency (19);<br>• US Centers for Disease Control (20);<br>• University of Miami Hospitals and Clinics, USA (28) |
| Non-governmental organisation | Global Challenges Research Fund Project (21) |
| International governmental organisation | European Centre for Disease Prevention and Control (29) |

### Policy objectives and role of modelling

Infectious disease modelling was done to cater various policy objectives. Most modelling efforts were done to guide operational planning and decision-making in the context of high uncertainty. Mathematical models were used in understanding transmission dynamics, and to forecast the scale of disease outbreak by providing both short- and long-term projections of confirmed cases, hospitalizations, and deaths, while aiding in healthcare resource management and allocation such as hospital bed occupancy and staffing needs (18, 20, 24, 26, 30–32) (15).

Many modelling approaches were also undertaken to assess the effects and impacts of various mitigation strategies and answer policy questions under different disease outbreak scenarios. For instance, during the COVID-19 pandemic, modelling determined the timeframe of the lifting of the state of emergency (SOE) in Tokyo while controlling infection and minimizing the economic loss (33); while the CoMo Consortium’s model was used to determine lockdown and school opening and closing strategies (16). In New York, the modelling aimed to meet the benchmark of the ‘HIV Ending the Epidemic Initiative’ under the most realistic conditions, with a focus on prioritizing men who have sex with men (22).

### Modelling approach

In 15 of the modelling efforts, practitioners developed and used compartmental models, i.e. models based on differential equations. 14 of the articles used Agent Based Models (e.g. (27)) or stochastic/statistical models (e.g. (34)). 10 studies mentioned more statistical approaches, such as using neural networks or Generalized Additive Models (GAM) (e.g. (19, 29, 32, 35)). 4 studies utilized ensemble approaches, synthesizing results from multiple models that included Agent based, Stochastic and compartmental models among others - often by different modelling teams (e.g. (20)). Note many articles considered multiple models and model types.

### Decision maker

Infectious disease modelling was used to inform decision makers across multiple governance levels. (Table 4). While some articles considered decision makers on one governance level, some considered multiple levels, e.g. in the Philippines (36) and South Africa (26).

**Table 4:**
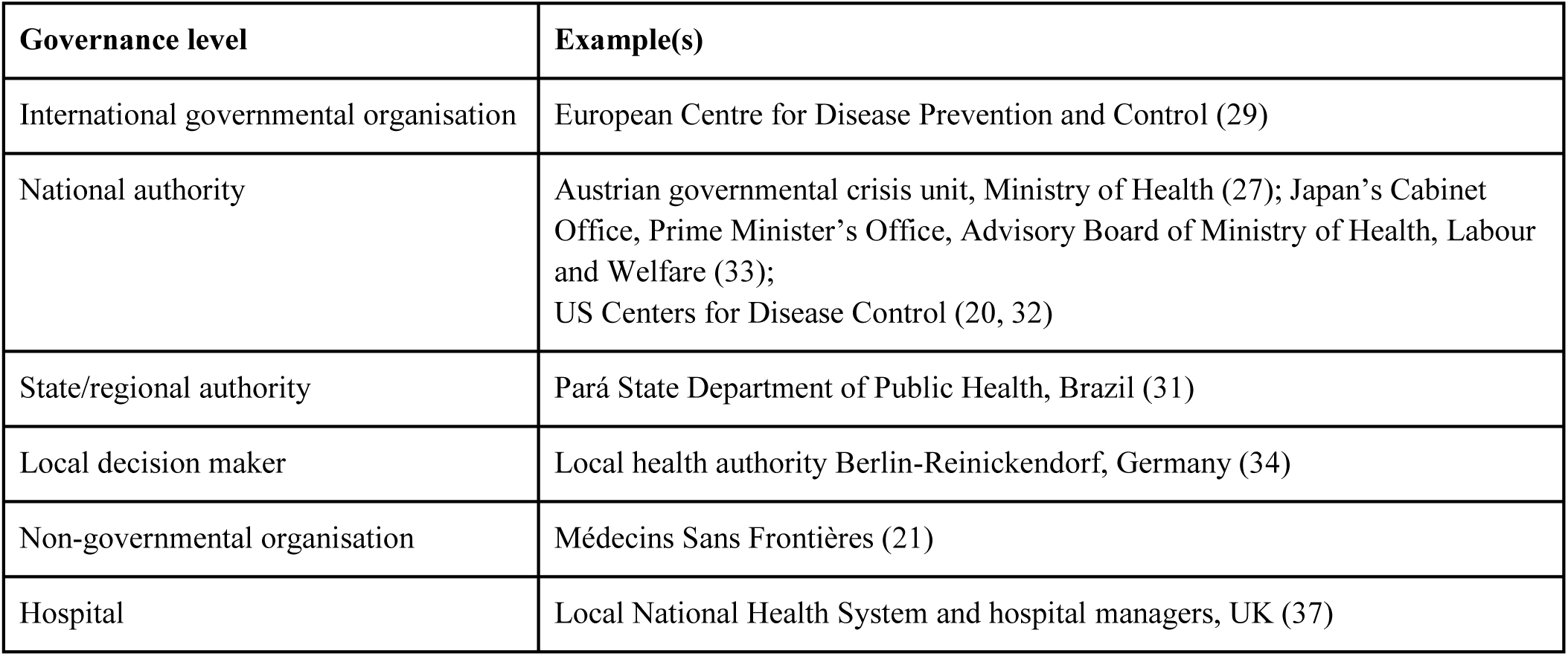
Infectious disease modelling was used to inform decision makers across multiple governance levels. Decision makers who used modelling to inform policy were not only from political institutions but also from e.g. hospitals.

| Governance level | Example(s) |
| --- | --- |
| International governmental organisation | European Centre for Disease Prevention and Control (29) |
| National authority | Austrian governmental crisis unit, Ministry of Health (27); Japan's Cabinet Office, Prime Minister's Office, Advisory Board of Ministry of Health, Labour and Welfare (33);<br>US Centers for Disease Control (20, 32) |
| State/regional authority | Pará State Department of Public Health, Brazil (31) |
| Local decision maker | Local health authority Berlin-Reinickendorf, Germany (34) |
| Non-governmental organisation | Médecins Sans Frontières (21) |
| Hospital | Local National Health System and hospital managers, UK (37) |

### Approaches to policy interaction

We identified various communication formats used by modellers to convey their findings to policymakers, other modellers, and other relevant professionals and organizations.

These communication approaches ranged from formalized, structured processes to more ad-hoc arrangements. Formal structures included established reporting templates (27), regular meeting schedules (e.g. (22)), and dedicated communication channels (e.g. (17)). Diverse communication methods were employed for these interactions, from daily reports (e.g. (28)) to weekly communications (e.g. (38)) and monthly meetings (e.g. (14)). The formats included written reports (e.g. (18)), face-to-face or virtual meetings (e.g. (39)), and formal presentations (e.g. (16)). Several initiatives established dedicated coordination teams or workstreams to manage the interface between modellers and decision-makers (e.g. (40)).

Hierarchical paths were frequently observed, where results were first presented to technical groups, then to advisory bodies, and finally to government through chief scientific advisors (e.g. (41)).

A notable theme across multiple studies was the emphasis on participatory and collaborative approaches to ensure effective knowledge translation. This was highlighted in The CoMo Consortium’s stepwise method to establish channels of communication between local policymakers and modellers, efforts by Médecins Sans Frontières and Canada’s bi-directional communication between modellers and decision- makers via multiple workshops to identify translational barriers (17, 21, 23). The adaptation of communication methods to different audiences was also common, with both technical reports and simplified briefings for broader audiences. Nigeria’s approach with regular evening calls between technical teams and policy task forces enabled rapid response to emerging questions (14). Public communication with several studies incorporating press releases, media engagement, and public-facing websites ensured broader dissemination of modelling insights (e.g. (33)).

### Stakeholder input

Stakeholder input refers to the contributions, feedback, and interactions of individuals or groups with an interest in the modelling process or those who may be impacted by its outcomes. The input was incorporated in various aspects of the project such as during the selection of variables, setting assumptions, and identifying critical questions to address in the models (16, 39, 42), with most studies aiming for collaborative improvement of the modelling process via expert input (e.g. (28, 32)).

15 out of 33 studies benefited from stakeholder input, where stakeholders were often professionals, including policy makers, cabinet officers, public health professional, clinicians, socioeconomic experts and modellers with little to no community level engagement. This input was considered in most studies after the modelling process to improve models but collaborative approaches were also seen with input being incorporated throughout the project via iterative processes and sector-specific expertise to refine models. Such efforts are exemplified in New Zealand’s two-round modelling approaches that allowed for clarification between teams and ’scenario sandpit’ discussions (40), where policymakers requested specific simulations, and the formation of specialized working groups in Nigeria (14).

Stakeholder engagement occurred through various structured feedback mechanisms like surveys and Plan- Do-Study-Act cycles implemented among clinicians, hospital staff, commissioners, and public health consultants by National Health Service teams (37, 38).

Direct communication channels with advisory groups and the media were established in multiple projects, including those in Canada with Indigenous communities through ’Community of Practice’ models (23). Additionally, international collaborations like the CoMo Consortium emphasized the role of local experts in guiding policy decisions and ensuring models reflected local contexts (17).

### Validity assessments of model results

Modellers tried to ensure the validity of their models and results through diverse quantitative and qualitative methods across studies. Quantitative model validation approaches included comparing forecasts against actual data (e.g. (19)), calibrating models on actual data (e.g. (36)) and conducting sensitivity analyses (e.g. (33)). Cross-validation between different modelling approaches was also observed (e.g. (28)) and some studies utilized ensemble approaches, benefiting from the fact that combined forecasts often outperformed individual models (e.g. (20)). Decision makers themselves (in North Carolina, USA) also appreciated comparisons between different models and teams, gaining confidence in the results if they confirmed each other and uncertainty if they diverged (15).

Validation frequently involved qualitative iterative processes, such as real-time assessment of forecast performance (33), allowing for continuous refinement of models. Several initiatives also implemented peer review mechanisms, exemplified by New Zealand’s rapid multi-disciplinary review process (40) and the use of multiple independent modelling groups (e.g. (42)).

### Facilitators and Barriers

The included publications reflected on various facilitators and barriers to informing policies and decisions. They can be roughly clustered around the themes ‘data infrastructure’, ‘institutional and collaborative structures’, ‘expertise and capacity’, and ‘communication and trust’ (Table 5).

**Table 5:** Reported facilitators and barriers for informing policies and decisions with infectious disease modelling. Note that we include examples for the individual facilitators and barriers where the facilitator/barrier is clearly highlighted. This does not exclude that other included articles also consider the respective facilitator/barrier.

| Theme | Facilitators | Barriers |
| --- | --- | --- |
| Data infrastructure | • Formal data-sharing agreements (e.g. (35))<br>• Public data availability (e.g. (41)) | • Quality issues and inconsistencies (e.g. (36))<br>• Reporting delays (e.g. (21))<br>• Integration challenges (e.g. (24))<br>• Limited access to granular data (e.g. (19)) |
| Institutional and collaborative structures | • Pre-existing networks (e.g. (32))<br>• Multi-stakeholder partnerships (e.g. (18))<br>• Funding (e.g. (18, 23)) | • Fragmented decision-making (e.g. (14))<br>• Limited funding (e.g. (14)) |
| Expertise and capacity | • Diverse expertise (e.g. (14))<br>• Professionals in decision making bodies to interpret models and their results (e.g. (15)) | • Limited national modelling capacity (e.g. (16))<br>• Knowledge gaps (e.g. (21))<br>• Insufficient consideration of human behaviour in models (e.g. (15)) |
| Communication and trust | • Participatory approaches (e.g. (17))<br>• Feedback loops (e.g. (17))<br>• Active engagement platforms (e.g. (18))<br>• Good visualisations (e.g. (15)) | • Challenges in conveying uncertainty (e.g. (42))<br>• Misinterpretation of results (e.g. (43))<br>• Trust building challenges (e.g. (16))<br>• Reliance on key individuals (e.g. (17))<br>• Confusion about differing results from multiple models (e.g. (15)) |

The modelling process was strongly facilitated by well-established institutional and collaborative frameworks. Multi-stakeholder partnerships involving experts from government, decision-making bodies, and academia led to improved predictive accuracy and efficient process execution according to e.g. the US FluSight Network organized by the CDC (18, 20). This pre-existing Influenza modelling network also laid the groundwork for the COVID-19 and Influenza Scenario Modelling Hub, which offered long-term operational support to inform outbreak response policy in the US (18, 32). Substantial funding through research grants from the CDC further facilitated these modelling initiatives.

In contrast, inadequate institutional and collaborative structures were identified as significant barriers to the modelling process and the translation of evidence into policy. For example in Nigeria, several systematic challenges were observed, including fragmented decision-making structures, and limited availability of public health funding to support modelling efforts (14).

13 of the included studies highlighted barriers in data infrastructure. These challenges encompassed issues such as data unavailability (e.g. (33)), occasional unreliability of data (e.g. (44)), reporting delays (e.g. (21)), the presence of data from multiple sources leading to inconsistencies and integration difficulties (e.g. (24)), and restricted access to granular data, which sometimes lacked key epidemiological parameters (e.g. (19)).

6 of the reviewed articles highlighted uncertainties in the projections due to the novel nature of the coronavirus and the use of varying parameters and assumptions by different modelling teams. This led to a discordant set of results presented to policymakers, fostering confusion, mistrust, and diminishing interest in the models among decision-makers, as illustrated by a study in the Eastern Mediterranean region (16). Keeling et al. also noted the challenge of effectively communicating such uncertainties in long-term projections to policymakers and the public (42). The results were frequently misinterpreted as definitive predictions in the media and decision-making contexts, which, in some cases, led to the potential dismissal of the modelling outcomes (43).

Local decision makers were especially concerned about translating model results based on national data and perspectives into their local context and expressed scepticism towards the ability of models to appropriately incorporate human behaviour (15). In their case, having internal professionals to interpret and translate the results was reported as helpful.

## DISCUSSION

In this scoping review, we aimed to get an overview of the infectious disease modelling literature that assesses its utility in informing policy for outbreak response. We included 33 peer-reviewed articles, none of which systematically assessed the utility of their modelling effort for policy. Instead, authors more subjectively reflected on their modelling effort(s) and highlighted key aspects they deemed relevant for utility. We focused on different aspects of the described modelling-to-policy pathways: the context of the outbreaks, whether the authors and initiatives stemmed from the modelling or policy field, the objectives of the policy and the role of modelling in achieving these objectives, the types of decision makers, how modelling informed policy, and the facilitators and barriers in the pathway.

Our analysis reveals that there are multiple processes through which modelling is able to inform policy. A key finding is that modellers perceive the modelling-to-policy process to be enhanced by well- established institutional structures and collaborative frameworks, particularly through multi-stakeholder partnerships between policymakers and researchers (14, 18, 32, 35, 42). These frameworks not only facilitated the rapid generation of scientific evidence but also promoted open communication and trust between policymakers and modellers, which was critical for the co-creation of policy-relevant evidence during the pandemic.

However, our review also identified several barriers that were thought to be hindering the utility of modelling for the decision-making process. The barriers to using infectious disease modelling in policymaking emerged across several key themes. Data challenges—unavailable, unreliable, or delayed— undermined the accuracy of models, while integration issues tangled the flow of information. Institutional hurdles, like fragmented decision-making, stifled progress, especially where limited funding further constrained efforts (14). Limited expertise and national modelling capacity, compounded with knowledge gaps regarding the disease, further deepened these setbacks. Communication between modellers and policymakers proved to be another stumbling block in the modelling-to-policy process as uncertainties in projections sowed confusion and eroded trust among policymakers (45). Misinterpretation of modelling results by the media only amplified this uncertainty and diminished confidence in the models, undermining their potential impact on policy decisions (43).

## Limitations

A limitation of our study is its eligibility criteria: To use the ‘remove duplication function’ of Ovid and thereby ensure a manageable scope of the review, we had to limit the number of retrieved articles in the search to below 6000. We achieved this by adapting our eligibility and search criteria. This involved focusing on only key pathogens and the exclusion of studies on cost-effectiveness and pharmaceutical interventions, studies looking only at single non-pharmaceutical interventions for COVID-19, as well as not-peer-reviewed publications. Yet, we acknowledge that in doing so, we may have created bias in our sample of papers and overlooked some valuable insights. For example, the substantial body of work from modellers reporting specific non-pharmaceutical interventions for COVID-19 may represent a different experience of the modelling to policy process or emphasise different facilitators and barriers.

We also acknowledge that much of the modelling for policy that shapes real world decision making eludes the academic record. This includes grey literature such as situational reporting, software tools, and other real-time outputs that may be too fluid for traditional academic reporting structures (46, 47). Further, our review may be affected by publication bias. Authors may not identify policy interfacing work as appropriate to report in academic publications, or may prefer not to report attempts to inform policy that were perceived as particularly challenging or unsuccessful. This potentially might mean that only more manageable barriers were represented in our analysis.

While a future systematic review could omit some of our restrictive criteria to capture a broader picture of the existing literature, we believe our approach has nevertheless captured key characteristics of the evidence body because other commentaries and viewpoints on the same topic support our own findings (1–3, 5, 48–53).

Owek et al. incorporated a similar scoping review, concentrating on COVID-19 in LMICs and used a different search strategy, including different search terms (9). Although we did not actively exclude LMIC settings from our analysis, more papers from HICs met our inclusion criteria. To ensure a more diverse overview, we therefore also included articles from Owek et al.’s scoping review if they matched our eligibility criteria. The comparison between our findings and those of the study focusing on LMICs shows many similar insights, emphasizing the robustness of our results: Both reviews highlight that the efficacy of policy modelling efforts seems to increase with pre-existing connections between modellers and policy makers, as well as open communication and trust between them. A key difference is that Owek et al. reported rivalry among modellers and officials’ dislike of research in LMICs as barriers to effective knowledge translation (9). These barriers were not found in our review.

The integration of scientific evidence into policy making is not a straightforward, linear process, but rather a complex and multifaceted pathway. Epidemiological modelling evidence related to infectious disease outbreaks represents only one of many factors that shape this process. To systematically evaluate evidence-informed policy making, various models and frameworks have been developed to organize existing knowledge in policy development (54). These frameworks, commonly utilized in health policy and systems research, include the Ottawa Model of Research Use (OMRU) (55), The Framework for Research Dissemination and Utilization (FRDU) (56) and The Canadian Health Services Research Foundation model (CHSRF) (57), among others. Notably, none of the articles reviewed in this study conducted a systematic evaluation of the utility of modelling outputs in policy translation. Instead, the assessments were subjective, mostly reflecting the perspectives of the authors of modelling studies on what they considered important for the utility of the process. However, many of the facilitators identified in the modelling-to-policy pathway closely align with the guiding principles of the aforementioned frameworks for evidence-informed policy making. For example, both the OMRU and FRDU frameworks emphasize the importance of involving multiple stakeholders at various levels of the healthcare system in the knowledge-to-policy translation, while the CHSRF advocates for the creation of communication channels among researchers, decision makers, and research funders to foster trust and facilitate the mutual exchange of knowledge. Such collaborative approaches, particularly for modelling-to-policy pathways, are also praised by others (3, 50, 51), which further highlights the importance of bringing scientists and decision makers together for modelling initiatives.

## Conclusion

In this scoping review, we investigated the role of mathematical modelling in shaping policy decisions during infectious disease outbreaks, and identified the factors that practitioners considered crucial for the effectiveness of this process. The initial objective of this review was to identify literature evaluating the utility of mathematical modelling in informing policy during infectious disease outbreaks, which could potentially inform a future systematic review. However, we did not find any studies that provided systematic assessments of the utility or evaluations of the pathways from modelling to policy. Together with other previous commentaries (3,) this lack highlights the need for tools and studies that systematically perform these assessments and evaluations. These evaluations could involve prospectively defining criteria in collaboration with decision makers that would enable retrospective evaluation once the modelling work has completed, but also more general reflections of modellers on model accuracy, utility and the relationship between the two. Modelling work to inform policy has the potential to do harm as well as save lives, and only through honest evaluation of past efforts will it be possible to come to general insights on how to maximise its positive impact on human health.

## Supporting information

Data extraction table

PRISMA-ScR checklist

## Data Availability

All data produced in the present work are contained in the manuscript and supplementary material.
The review protocol can be accessed at https://osf.io/cd4qa/

## ACKNOWLEDGMENTS

We thank Francisco Pozo Martin for his guidance on conducting a literature review. We are grateful to Liza Hadley and Paula Christen for their feedback on our results.

## AUTHOR CONTRIBUTIONS

**Conceptualization:** SAM, JH, JF, SF, ENI

**Data curation:** KH, SAM, DR, AT, ENI, KS

**Formal analysis:** DR, AT, ENI

**Funding acquisition:** JH, SF, JF

**Investigation:** DR, AT, ENI

**Methodology:** SAM, DR, AT, ENI, KS, SF, PCB

**Project administration:** DR, AT, ENI, SAM

**Supervision:** SF, KS, JF, JH, ENI

**Validation:** DR, AT, ENI, SF, JF, KS

**Visualization:** DR, AT, ENI

**Writing – original draft:** DR, AT, ENI

**Writing – review & editing**: DR, AT, ENI, SF, JF, KR, SAM, KS

## REFERENCES

1. James LP, Salomon JA, Buckee CO, Menzies NA. The Use and Misuse of Mathematical Modeling for Infectious Disease Policymaking: Lessons for the COVID-19 Pandemic. Medical Decision Making. 2021;41(4):379–85.

2. Hadley L, Rich C, Tasker A, Restif O, Funk S. How does policy modelling work in practice? A global analysis on the use of modelling in Covid-19 decision-making. medRxiv. 2024.

3. Jit M, Ainslie K, Althaus C, Caetano C, Colizza V, Paolotti D, et al. Reflections On Epidemiological Modeling To Inform Policy During The COVID-19 Pandemic In Western Europe, 2020–23. Health Affairs. 2023;42(12):1630-6.

4. Lofgren ET, Halloran ME, Rivers CM, Drake JM, Porco TC, Lewis B, et al. Mathematical models: A key tool for outbreak response. Proceedings of the National Academy of Sciences. 2014;111(51):18095–6.

5. Le Rutte EA, Shattock AJ, Zhao C, Jagadesh S, Balać M, Müller SA, et al. A case for ongoing structural support to maximise infectious disease modelling efficiency for future public health emergencies: A modelling perspective. Epidemics. 2024;46:100734.

6. How modelling can better support public health policy making: the Lancet Commission on Strengthening the Use of Epidemiological Modelling of Emerging and Pandemic Infectious Diseases. Elsevier; 2024. p. 789–91.

7. Salajan A, Tsolova S, Ciotti M, Suk JE. To what extent does evidence support decision making during infectious disease outbreaks? A scoping literature review. Evidence & Policy. 2020;16(3):453–75.

8. Elsland Sv, Christen P. Advanced analytics to policy decision making in Europe: a systematic review on COVID-19 knowledge translation. PROSPERO2024.

9. Owek CJ, Guleid FH, Maluni J, Jepkosgei J, Were VO, Sim SY, et al. Lessons learned from COVID-19 modelling efforts for policy decision-making in lower- and middle-income countries. BMJ Global Health. 2024;9(11):e015247.

10. Müller S. Assessing the utility of infectious disease modelling in informing policy for outbreak response: a rapid review. OSF; 2024.

11. Tricco AC, Lillie E, Zarin W, O’Brien KK, Colquhoun H, Levac D, et al. PRISMA Extension for Scoping Reviews (PRISMA-ScR): Checklist and Explanation. https://doiorg/107326/M18-0850. 2018.

12. Pollock D, Peters MDJ, Khalil H, McInerney P, Alexander L, Tricco AC, et al. Recommendations for the extraction, analysis, and presentation of results in scoping reviews. JBI evidence synthesis. 2023;21(3).

13. Munn Z, Peters MDJ, Stern C, Tufanaru C, McArthur A, Aromataris E. Systematic review or scoping review? Guidance for authors when choosing between a systematic or scoping review approach. BMC Medical Research Methodology. 2018;18(1):143.

14. Abubakar I, Dalglish SL, Ihekweazu CA, Bolu O, Aliyu SH. Lessons from co-production of evidence and policy in Nigeria’s COVID-19 response. 2021.

15. Johnson K, Biddell CB, Hassmiller Lich K, Swann J, Delamater P, Mayorga M, et al. Use of Modeling to Inform Decision Making in North Carolina during the COVID-19 Pandemic: A Qualitative Study. MDM Policy & Practice. 2022;7(2):23814683221116362.

16. Adib K, Hancock PA, Rahimli A, Mugisa B, Abdulrazeq F, Aguas R, et al. A participatory modelling approach for investigating the spread of COVID-19 in countries of the Eastern Mediterranean Region to support public health decision-making. BMJ Global Health. 2021;6(3):e005207.

17. Aguas R, White L, Hupert N, Shretta R, Pan-Ngum W, Celhay O, et al. Modelling the COVID-19 pandemic in context: an international participatory approach. BMJ Global Health. 2020;5(12):e003126.

18. Loo SL, Howerton E, Contamin L, Smith CP, Borchering RK, Mullany LC, et al. The US COVID-19 and Influenza Scenario Modeling Hubs: Delivering long-term projections to guide policy. Epidemics. 2024;46:100738.

19. Mellor J, Christie R, Overton CE, Paton RS, Leslie R, Tang M, et al. Forecasting influenza hospital admissions within English sub-regions using hierarchical generalised additive models. Communications medicine. 2023;3(1):190.

20. Reich NG, McGowan CJ, Yamana TK, Tushar A, Ray EL, Osthus D, et al. Accuracy of real-time multi- model ensemble forecasts for seasonal influenza in the U.S. PLoS Computational Biology. 2019;15(11):e1007486.

21. Finger F, Funk S, White K, Siddiqui MR, Edmunds WJ, Kucharski AJ. Real-time analysis of the diphtheria outbreak in forcibly displaced Myanmar nationals in Bangladesh. BMC Medicine. 2019;17(1):58.

22. Martin EG, MacDonald RH, Gordon DE, Swain CA, O’Donnell T, Helmeset J, et al. Simulating the End of AIDS in New York: Using Participatory Dynamic Modeling to Improve Implementation of the Ending the Epidemic Initiative. Public Health Reports. 2020;135(1_suppl):158S-71S.

23. Tariq M, Haworth-Brockman M, Moghadas SM. Ten years of Pan-InfORM: modelling research for public health in Canada. AIMS public health. 2021;8(2):265–74.

24. Steinberg DM, Balicer RD, Benjamini Y, De-Leon H, Gazit D, Rossman H, et al. The role of models in the covid-19 pandemic. Israel Journal of Health Policy Research. 2022;11(1):36.

25. Sanchez F, Calvo JG, Mery G, Garcia YE, Vasquez P, Barboza LA, et al. A multilayer network model of Covid-19: Implications in public health policy in Costa Rica. Epidemics. 2022;39:100577.

26. Silal SP, Groome MJ, Govender N, Pulliam JRC, Ramadan OP, Puren A, et al. Leveraging epidemiology as a decision support tool during the COVID-19 epidemic in South Africa. South African Medical Journal. 2022;112(5b):361–5.

27. Bicher M, Zuba M, Rainer L, Bachner F, Rippinger C, Ostermann H, et al. Supporting COVID-19 policy- making with a predictive epidemiological multi-model warning system. Communications medicine. 2022;2(1):157.

28. Warde PR, Patel S, Ferreira T, Gershengorn H, Bhatia MC, Parekh D, et al. Linking prediction models to government ordinances to support hospital operations during the COVID-19 pandemic. BMJ health & care informatics. 2021;28(1).

29. Sherratt K, Srivastava A, Ainslie K, Singh DE, Cublier A, Marinescu MC, et al. Characterising information gains and losses when collecting multiple epidemic model outputs. Epidemics. 2024;47:100765.

30. Morozova O, Li ZR, Crawford FW. One year of modeling and forecasting COVID-19 transmission to support policymakers in Connecticut. Scientific reports. 2021;11(1):20271.

31. Souza GNd, Jr., Braga MdB, Rodrigues LLS, Fernandes RdS, Ramos RTJ, Carneiro AR, et al. COVID- PA Bulletin: reports on artificial intelligence-based forecasting in coping with COVID-19 pandemic in the state of Para, Brazil. Boletim COVID-PA: relatos sobre projecoes baseadas em inteligencia artificial no enfrentamento da pandemia de COVID-19 no estado do Para. 2021;30(4):e2021098.

32. Shea K, Borchering RK, Probert WJM, Howerton E, Bogich TL, Li SL, et al. Multiple models for outbreak decision support in the face of uncertainty. Proceedings of the National Academy of Sciences of the United States of America. 2023;120(118):e2207537120.

33. Fujii D, Nakata T. COVID-19 and output in Japan. Japanese economic review (Oxford, England). 2021;72(4):609–50.

34. Jackle S, Alpers R, Kuhne L, Schumacher J, Geisler B, Westphal M. EsteR - A Digital Toolkit for COVID-19 Decision Support in Local Health Authorities. Studies in health technology and informatics. 2022;296:17–24.

35. Pellis L, Scarabel F, Stage HB, Overton CE, Chappell LHK, Fearon E, et al. Challenges in control of COVID-19: short doubling time and long delay to effect of interventions. Philosophical transactions of the Royal Society of London Series B, Biological sciences. 2021;376(1829):20200264.

36. de Lara-Tuprio E, Estadilla CDS, Macalalag JMR, Teng TR, Uyheng J, Espina KE, et al. Policy-driven mathematical modeling for COVID-19 pandemic response in the Philippines. Epidemics. 2022;40:100599.

37. Irvine N, Anderson G, Sinha C, McCabe H, van der Meer R. Collaborative critical care prediction and resource planning during the COVID-19 pandemic using computer simulation modelling: Future urgent planning lessons. Future Healthcare Journal. 2021;8(2):E317–E21.

38. George A, Lacey P, Badrinath P, Gray A, Turner P, Harwood C, et al. Planning for healthcare services during the COVID-19 pandemic in the Southeast of England: A system dynamics modelling approach. BMJ Open. 2023;13(12):e072975.

39. Howerton E, Contamin L, Mullany LC, Qin M, Reich NG, Bents S, et al. Evaluation of the US COVID- 19 Scenario Modeling Hub for informing pandemic response under uncertainty. Nat Commun. 2023;14(7260):1– 15.

40. Hendy S, Steyn N, James A, Plank MJ, Hannah K, Binny RN, et al. Mathematical modelling to inform New Zealand’s COVID-19 response. Journal of the Royal Society of New Zealand. 2021.

41. Sherratt K, Abbott S, Meakin SR, Hellewell J, Munday JD, Bosse N, et al. Exploring surveillance data biases when estimating the reproduction number: with insights into subpopulation transmission of COVID-19 in England. Philosophical transactions of the Royal Society of London Series B, Biological sciences. 2021;376(1829):20200283.

42. Keeling MJ, Dyson L, Tildesley MJ, Hill EM, Moore S. Comparison of the 2021 COVID-19 roadmap projections against public health data in England. Nature Communications. 2022;13(1):4924.

43. Mccaw JM, Plank MJ. THE ROLE OF THE MATHEMATICAL SCIENCES IN SUPPORTING THE COVID-19 RESPONSE IN AUSTRALIA AND NEW ZEALAND. ANZIAM J. 2022;64(4):315–37.

44. Turk PJ, Anderson WE, Burns RJ, Chou SH, Dobbs TE, Kearns JT, et al. A regionally tailored epidemiological forecast and monitoring program to guide a healthcare system in the COVID-19 pandemic. Journal of Infection and Public Health. 2024;17(6):1125–33.

45. McCabe R, Donnelly CA. Disease transmission and control modelling at the science–policy interface. Interface Focus. 2021;11(6):20210013.

46. Kucharski AJ, Funk S, Eggo RM. The COVID-19 response illustrates that traditional academic reward structures and metrics do not reflect crucial contributions to modern science. PLOS Biology. 2020;18(10):e3000913.

47. Sherratt K, Carnegie A, Kucharski A, Cori A, Pearson C, Jarvis C, et al. Improving modelling for epidemic responses: reflections from members of the UK infectious disease modelling community on their experiences during the COVID-19 pandemic [version 1; peer review: 2 approved]. Wellcome Open Research. 2024;9(12).

48. Adiga A, Dubhashi D, Lewis B, Marathe M, Venkatramanan S, Vullikanti A. Mathematical Models for COVID-19 Pandemic: A Comparative Analysis. Journal of the Indian Institute of Science. 2020;100(4):793–807.

49. Becker AD, Grantz KH, Hegde ST, Bérubé S, Cummings DAT, Wesolowski A. Development and dissemination of infectious disease dynamic transmission models during the COVID-19 pandemic: what can we learn from other pathogens and how can we move forward? The Lancet Digital Health. 2021;3(1):e41–e50.

50. Dial NJ, Croft SL, Chapman LAC, Terris-Prestholt F, Medley GF. Challenges of using modelling evidence in the visceral leishmaniasis elimination programme in India. PLOS Global Public Health. 2022;2(11):e0001049.

51. Hadley L, Challenor P, Dent C, Isham V, Mollison D, Robertson DA, et al. Challenges on the interaction of models and policy for pandemic control. Epidemics. 2021;37:100499.

52. IVIR-AC. Meeting of the Immunization and Vaccine-related Implementation Research Advisory Committee (IVIR-AC), September 2023. World Health Organization. 2024:1–12.

53. van Elsland S, L. , Christen P. Political decision-makers and mathematical modellers of infectious disease outbreaks: the sweet spot for engagement. BMJ Global Health. 2024;9(9):e015155.

54. Jabali SH, Yazdani S, Pourasghari H, Maleki M. From bench to policy: a critical analysis of models for evidence-informed policymaking in healthcare. Frontiers in Public Health. 2024;12.

55. Logan J, Graham ID. Toward a Comprehensive Interdisciplinary Model of Health Care Research Use. Science Communication. 1998;20(2):227–46.

56. Dobbins M, Ciliska D, Cockerill R, Barnsley J, DiCenso A. A Framework for the Dissemination and Utilization of Research for Health-Care Policy and Practice. Worldviews on Evidence-based Nursing presents the archives of Online Journal of Knowledge Synthesis for Nursing. 2002;E9(1):149-60.

57. Medicine Io. An Integrated Framework for Assessing the Value of Community-Based Prevention. Washington (DC): National Academies Press (US)Copyright 2012 by the National Academy of Sciences. All rights reserved.; 2012.

