## Supplementary Information for "The utility of infectious disease modelling in informing policy for outbreak response: a scoping review"

**Methods: Details of the search strategy**

As mentioned in the main text, the literature search was conducted via Ovid. The final search from June 5 2024 is depicted in Supplementary Figure 1. The rationales behind the components of the total search string are described in Supplementary Table 1.

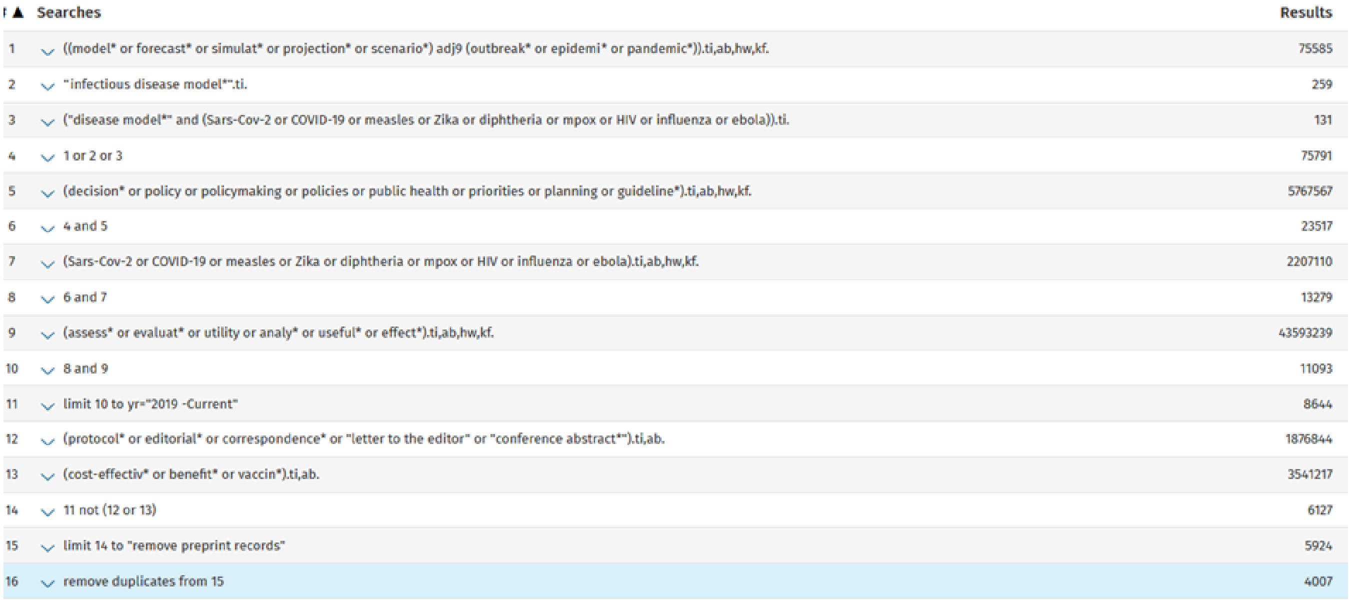

| # | Searches | Results |
|---|---|---|
| 1 | ((model* or forecast* or simulat* or projection* or scenario*) adj9 (outbreak* or epidemi* or pandemic*)).ti,ab,hw,kf. | 75585 |
| 2 | "infectious disease model*".ti. | 259 |
| 3 | ("disease model*" and (Sars-Cov-2 or COVID-19 or measles or Zika or diphtheria or mpox or HIV or influenza or ebola)).ti. | 131 |
| 4 | 1 or 2 or 3 | 75791 |
| 5 | (decision* or policy or policymaking or policies or public health or priorities or planning or guideline*).ti,ab,hw,kf. | 5767567 |
| 6 | 4 and 5 | 23517 |
| 7 | (Sars-Cov-2 or COVID-19 or measles or Zika or diphtheria or mpox or HIV or influenza or ebola).ti,ab,hw,kf. | 2207110 |
| 8 | 6 and 7 | 13279 |
| 9 | (assess* or evaluat* or utility or analy* or useful* or effect*).ti,ab,hw,kf. | 43593239 |
| 10 | 8 and 9 | 11093 |
| 11 | limit 10 to yr="2019 -Current" | 8644 |
| 12 | (protocol* or editorial* or correspondence* or "letter to the editor" or "conference abstract*").ti,ab. | 1876844 |
| 13 | (cost-effectiv* or benefit* or vaccin*).ti,ab. | 3541217 |
| 14 | 11 not (12 or 13) | 6127 |
| 15 | limit 14 to "remove preprint records" | 5924 |
| 16 | remove duplicates from 15 | 4007 |

*Supplementary Figure 1: Screenshot of final search strategy and query on Ovid.*

| **Lines in search strategy** | **Rationale** |
|---|---|
| 1-4 | Include all publications that are related to disease models to consider the outbreak model inclusion criterion |
| 5-6 | Require a connection to policy to consider the outbreak model inclusion criterion |
| 7-8 | Fulfill the inclusion criterion considering certain diseases |
| 9-10 | Fulfill policy evaluation inclusion criterion |
| 11 | Fulfill time inclusion criterion |
| 12+15 | Fulfill peer-review inclusion criterion |

| | |
|---|---|
| 13 | Fulfill pharmaceutical intervention and cost-effectiveness exclusion criteria |
| 14 | (See the above the rows) |
| 16 | Remove duplicates |

*Supplementary Table 1: Rationales for the search strategy and query on Ovid.*

**Results: Additional characteristics of the included articles**

The majority of the studies included were published in public health journals with the most number of articles being from the ‘British Medical Journal’ followed by the journal ‘Epidemics’.

Within the considered time frame from 2019-2024, most publications were from 2021 (Supplementary Figure 2). This is in line with the fact that most articles considered the COVID-19 pandemic.

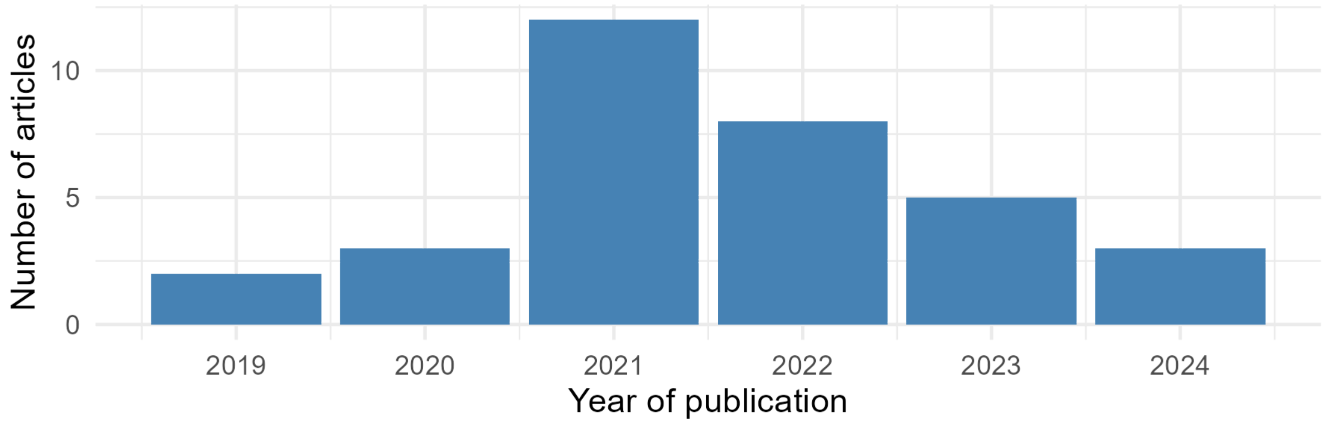


*Supplementary Figure 2: Most included publications are from the first years of the COVID-19 pandemic.*

The modelling efforts that are described in the included articles consider a variety of target populations within a geographical area (Supplementary Figure 3). However, most of them consider the total population of a country or of a region in a country.

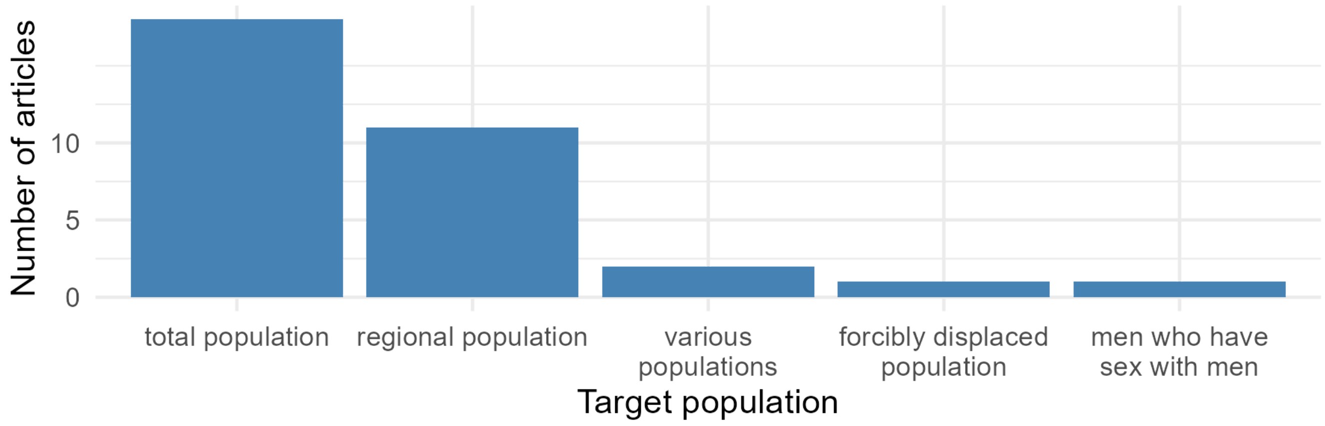


*Supplementary Figure 3: In their modelling efforts, included articles mostly consider the total population of a country or of a region within.*

Even though the search went beyond COVID-19 related articles, most included articles consider COVID-19 (Supplementary Figure 4).

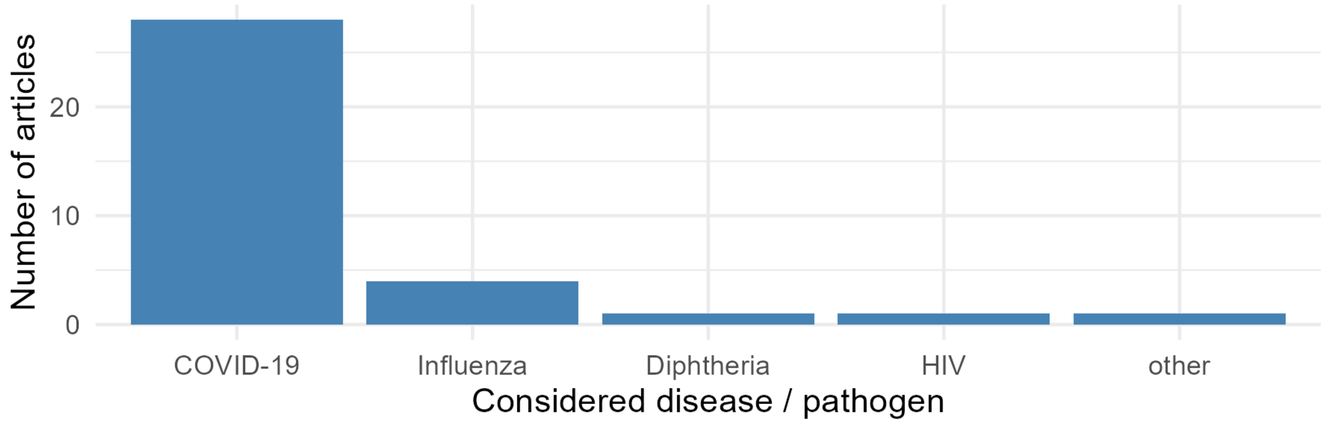


*Supplementary Figure 4:* ***Included articles mostly consider COVID-19.*** *Note that some articles consider multiple diseases/pathogens.*
